# Utilizing Large Language Models for Enhanced Clinical Trial Matching: A Study on Automation in Patient Screening

**DOI:** 10.1101/2024.04.10.24305571

**Authors:** Jacob Beattie, Sarah Neufeld, Daniel Yang, Christian Chukwuma, Ahmed Gul, Neil Desai, Steve Jiang, Michael Dohopolski

## Abstract

**Background:** Clinical trial matching, essential for advancing medical research, involves detailed screening of potential participants to ensure alignment with specific trial requirements. Research staff face challenges due to the high volume of eligible patients and the complexity of varying eligibility criteria. The traditional manual process, both time-consuming and error-prone, often leads to missed opportunities. Utilizing Artificial Intelligence (AI) and Natural Language Processing (NLP) can significantly enhance the accuracy and efficiency of this process through automated patient screening against established criteria.

**Methods:** Utilizing data from the National NLP Clinical Challenges (n2c2) 2018 Challenge, we utilized 202 longitudinal patient records. These records were annotated by medical professionals and evaluated against 13 selection criteria encompassing various health assessments. Our approach involved embedding medical documents into a vector database to determine relevant document sections, then using a large language model (GPT-3.5 Turbo and GPT-4 OpenAI API) in tandem with structured and chain-of-thought prompting techniques for systematic document assessment against the criteria. Misclassified criteria were also examined to identify classification challenges.

**Results:** This study achieved an accuracy of 0.81, sensitivity of 0.80, specificity of 0.82, and a micro F1 score of 0.79 using GPT-3.5 Turbo, and an accuracy of 0.87, sensitivity of 0.85, specificity of 0.89, and micro F1 score of 0.86 using GPT-4 Turbo. Notably, some criteria in the ground truth appeared mislabeled, an issue we couldn’t explore further due to insufficient label generation guidelines on the website.

**Conclusion:** Our findings underscore the significant potential of AI and NLP technologies, including large language models, in the clinical trial matching process. The study demonstrated strong capabilities in identifying eligible patients and minimizing false inclusions. Such automated systems promise to greatly alleviate the workload of research staff and improve clinical trial enrollment, thus accelerating the process and enhancing the overall feasibility of clinical research.

## 1. Introduction

Clinical trials are essential for advancing medical knowledge and introducing new treatment paradigms. However, many eligible patients miss out on participating in these trials due to a lack of discussion with their treatment teams, a problem rooted in challenges, such as resource scarcity for patient screening [1, 5]. Screening is often manual and inefficient, consuming up to 45 minutes per patient and contributing to a 3-9 hour process for a single enrollment [12]. Physicians, nurses, and research staff highlight time constraints and inadequate support as key obstacles [4, 6]. This situation contributes to low enrollment rates, leading to the premature closure of studies and limiting the scope of their findings [13]. The advent of automated patient screening technologies offers a promising solution by facilitating the identification of potential participants and alleviating the workload on research teams, aiming to enhance patient recruitment efficiency and trial outcomes.

Efforts to automate participant selection have shown promise. Applications that perform automatic matching based on genetic biomarkers have seen relative success, and this workflow was able to perform in real-time with improved accuracy [3]. Natural language processing (NLP) has also been used to analyze unstructured data sources like clinical notes. In combination with additional structured data, results from 2015 showed that these techniques could significantly reduce the patient-trial matching workload [8]. However, these approaches have substantial limitations. Methods relying on information extraction techniques fail to interpret semantic relations correctly [8], and several of these older, existing methods that process free text still require manual preprocessing from domain experts [3]. These barriers may prevent a large-scale implementation across a hospital system as they fail to work across all types of criteria or do not fully address clinical research staff shortages.

In recent advancements, NLP techniques and large language models (LLMs) have significantly evolved, showing great potential in transforming clinical trial eligibility screening. The screening process, inherently reliant on interpreting extensive unstructured text, finds a promising solution in NLP and LLMs due to their advanced reasoning and semantic understanding capabilities [19]. Despite their promise, the comprehensive application and in-depth evaluation of LLMs, including GPT-3.5 Turbo [10], GPT-4 [11], and Llama2 [15], for clinical trial patient screening have been sparse [21].

Our research aims to bridge this gap by employing state-of-the-art LLMs to directly analyze unstructured clinical data, thereby accurately determining patient eligibility for clinical trials. Through this endeavor, we aspire to significantly enhance the efficiency and accuracy of identifying eligible trial participants.

## 2 Methods

### 2.1 Data

We utilized the Harvard University National NLP Clinical Challenges (n2c2) 2018 cohort selection challenge dataset. This dataset comprises 288 longitudinal patient records and information regarding 13 selection criteria. These criteria include drug abuse, alcohol abuse, English proficiency, decisionmaking capacity, history of intra-abdominal surgery, major diabetes-related complications, advanced cardiovascular disease, dietary supplement intake in the past two months (excluding Vitamin D), diagnosis of ketoacidosis in the past year, aspirin use for myocardial infarction prevention, HbA1C values outside the 6.5%-9.5% range, abnormal creatinine levels, and myocardial infarction occurrence within the past six months. Each patient record was annotated by two medical experts, with any discrepancies resolved through adjudication by a researcher in consultation with a physician. [14]. Of the 288 patient records in the n2c2 2018 challenge dataset, 202 were made publicly available for training. We utilized 20 patient records for prompt engineering, while the 182 remaining records were reserved for testing.

### 2.2 Indexing and Document Transformation

To analyze patient records against specific criteria, we employed GPT-3.5 Turbo and GPT-4, mindful of their context length limits of 16,385 and 32,768 tokens, respectively. This limitation necessitated the careful selection of the most pertinent segments from a patient’s Electronic Health Record (EHR), as the full EHR could not be directly processed.

**Figure 1:**
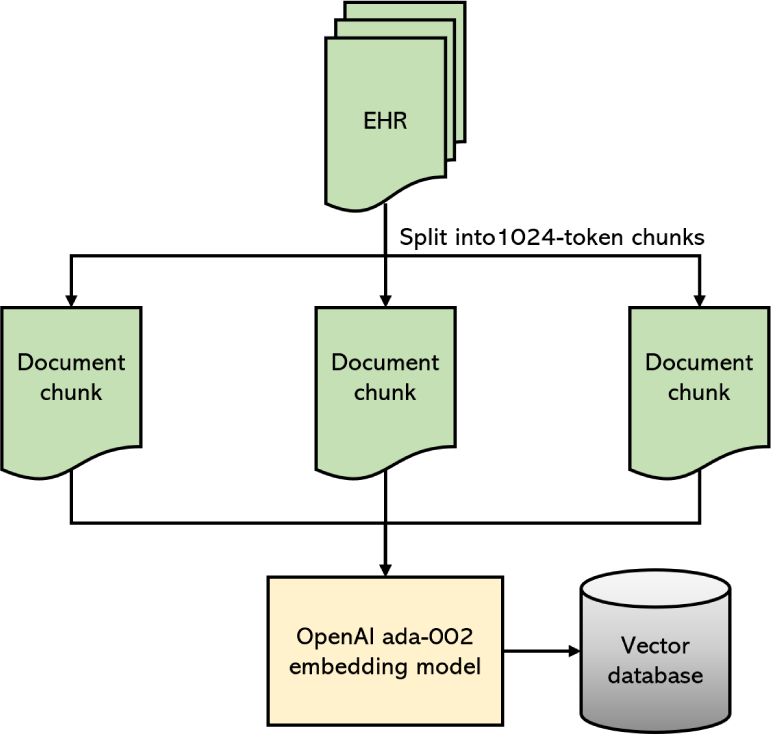
_Data indexing process._

**Figure 2:**
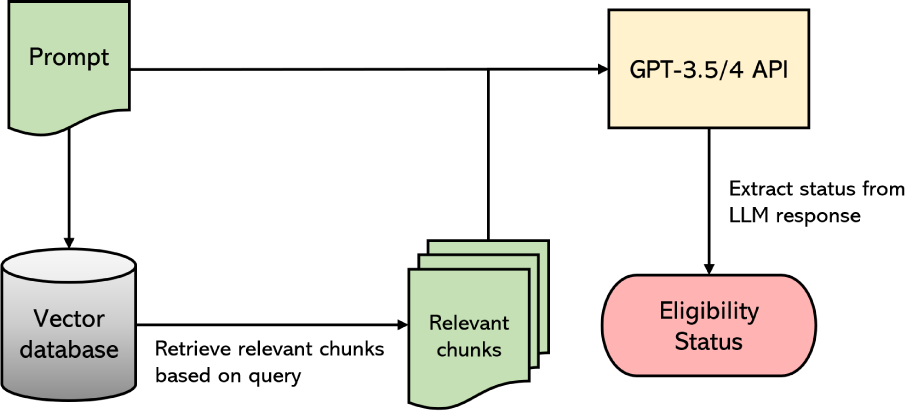
Eligibility screening process for a single patient on a single criterion.

Initially, we transformed the patient EHR documents into a format suitable for querying. Using LlamaIndex, documents were indexed by first partitioning each into 1024-token sections with a 10% overlap to ensure contextual continuity—a process known as chunking. These chunks were then embedded into a vector database using OpenAI’s ada-002 embedding model, creating vector representations for similarity search and retrieval. The embedded chunks and their original text were stored as key-value pairs in a database, facilitating the identification of the top-*k* chunks most semantically similar to a given query’s embedding.

### 2.3 Retrieval-Augmented Generation (RAG) with LlamaIndex

LlamaIndex enhanced GPT-3.5 Turbo and GPT-4’s accuracy by providing targeted context (EHR text chunks) through Retrieval-Augmented Generation (RAG). We created a retrieval system that selects the top-*k* relevant text passages for a given query. For each criterion, we set *k* = 5 and used the LLM prompt as the document retrieval search query. This process yielded the five most relevant chunks of the patient’s EHR.

### 2.4 Criteria Eligibility Analysis

The relevant EHR portions obtained were appended to the prompt, and GPT-3.5 Turbo was then applied to predict the criteria eligibility status. After processing all five relevant chunks, the final response was saved for analysis. This iterative process of retrieving context and applying the LLM to the prompt/context combination was repeated for each criterion for each patient, culminating in a comprehensive automated eligibility screening depicted in **Figures 1** and **2**.

### 2.5 Prompt Engineering

In our approach, we leveraged the principles of zero-shot learning with GPT-3.5 Turbo and GPT-4, applying a dynamic prompting strategy to evaluate patient eligibility for clinical trials from the n2c2 dataset[20, 7]. This strategy utilized a general template, customized with criterion-specific expert guidance and relevant excerpts from patient records to generate tailored prompts for each case (**Figure 3**). This method combines the zero-shot learning capability of making classifications without direct prior examples of the task with the adaptability of customized prompts, ensuring accuracy and relevance in a clinical context.

The prompt engineering process began with two foundational templates outlining the structure of the inquiry to the LLM: one focused on producing a structured JSON output, and the other focused on producing chain-of-thought (CoT) reasoning. This template was enriched with expert-generated tips to add in targeted patient data retrieval, creating a unique prompt for each eligibility criterion.

**Figure 3:**
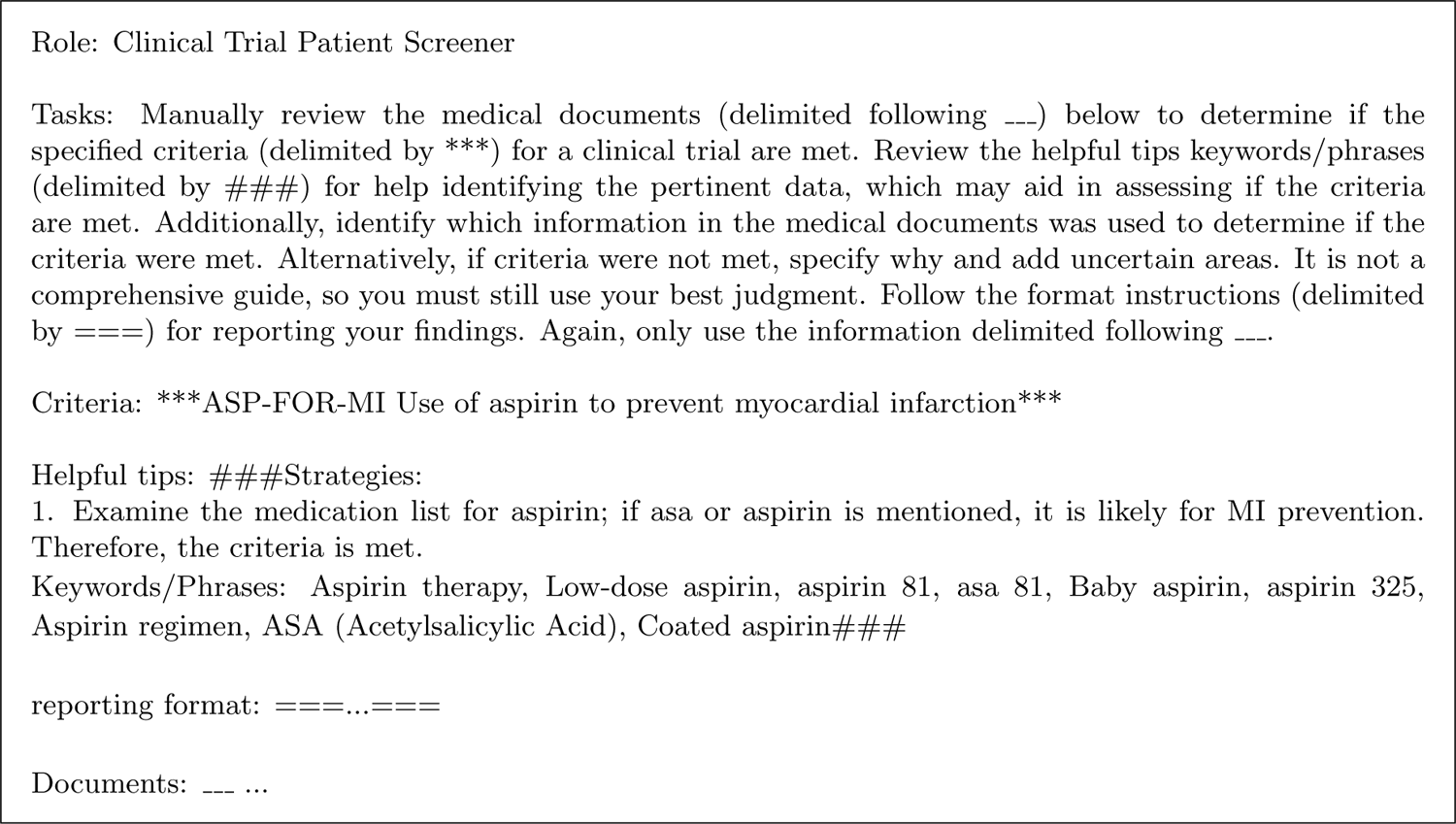
Example prompt for producing JSON output using manually created expert guidance

To fully automate the screening process, we also created a set of LLM-generated prompts to provide guidance; we used GPT-4 to create criteria descriptions and instructions. These LLM-generated tips consist of vocabulary commonly associated with the criteria being met or not met, emulating rule-based approaches used by the winning team of the original n2c2 challenge [9]. Full prompts for both JSON and CoT output, as well as examples of manual and LLM-created expert tips, can be found in **Appendix A**.

Through iterative testing with a subset of 20 patients, we identified and addressed discrepancies between the LLM’s outputs and the ground truth, refining the manually-created expert guidance and prompt templates. This iterative cycle of customization and evaluation continued until we achieved significant accuracy and micro F1 improvements, defined as 0.85, as this was the competitive performance in the n2c2 competition. The two prompting structures were applied to the n2c2 dataset, and their performance was compared. These optimized prompts, embodying the refined integration of expert guidance and patient-specific information, were subsequently applied across the entire dataset. This approach not only harnessed the power of zero-shot learning for efficient patient screening but also enhanced it through tailored prompts, striking a balance between the flexibility of zero-shot learning and the precision needed for clinical applicability.

### 2.6 Analyses

We assessed our model’s performance using accuracy, recall, precision, specificity, and micro F1 metrics across all criteria and patients. Individual criteria were analyzed to highlight specific classification challenges. Our results were benchmarked against the leading teams from the 2018 n2c2 competition to evaluate our standing.

In understanding our model’s limitations, we analyzed misclassifications (false negatives and false positives) by reviewing the LLM’s generated rationales and cross-referencing them with patient charts. This process aimed to identify common error patterns and underlying reasons for inaccuracies, informing future model improvements.

## 3 Results

In our testing encompassing 2366 criteria from 182 patient EHRs from the n2c2 dataset using GPT-

3.5 Turbo, we achieved an overall accuracy of 0.81, sensitivity of 0.80, specificity of 0.82, and micro F1 of 0.79. For GPT-3.5 Turbo, the best-performing approach utilized structured JSON output and manually created expert guidance. Additionally, we tested our approach on a smaller subset of data (40 patients) due to cost limitations, using GPT-4. Here, we observed an accuracy of 0.87, sensitivity of 0.85, specificity of 0.89, and micro F1 score of 0.86. Here, the best-performing approach again utilized structured JSON output and manually created expert guidance. This model’s performance across each criterion is listed in table 3. Our approach processes a single patient for all included criteria in 1-5 minutes. Variability was due to GPT3.5 and 4 resource allocations during experimentation.

**Table 1:** GPT-3.5 Turbo results when applied to test data (182 patients)

| Prompting Format | Accuracy | Sensitivity | Specificity | Precision | Micro F1 |
| --- | --- | --- | --- | --- | --- |
| Structured output + manual expert guidance | 0.8081 | 0.7988 | 0.8154 | 0.7721 | 0.7852 |
| Structured output + LLM-generated expert guidance | 0.6593 | 0.6006 | 0.7054 | 0.6148 | 0.6076 |
| CoT output + manual expert guidance | 0.7747 | 0.6064 | 0.9066 | 0.8355 | 0.7027 |
| CoT output + LLM-generated expert guidance | 0.6416 | 0.3367 | 0.8802 | 0.6876 | 0.4522 |

**Table 2:** GPT-4 results when applied to a subset of test data (40 patients)

| Prompting Format | Accuracy | Sensitivity | Specificity | Precision | Micro F1 |
| --- | --- | --- | --- | --- | --- |
| Structured output + manual expert guidance | 0.8692 | 0.8449 | 0.8909 | 0.8734 | 0.8589 |
| Structured output + LLM-generated expert guidance | 0.7712 | 0.6490 | 0.8800 | 0.8281 | 0.7277 |
| CoT output + manual expert guidance | 0.8558 | 0.8694 | 0.8436 | 0.8320 | 0.8503 |
| CoT output + LLM-generated expert guidance | 0.8519 | 0.8571 | 0.8473 | 0.8333 | 0.8451 |

### 3.1 Failure Analysis

We performed a thorough failure analysis of the best-performing approach, GPT-4, with structured JSON output and manually created expert guidance. The analysis pinpointed four criteria— DIETSUPP-2MOS, ADVANCED-CAD, and MI-6MOS —with significantly low accuracy rates, with MI-6MOS exhibiting the poorest performance at an accuracy of only 0.65. For MI-6MOS, this suboptimal performance primarily resulted from the LLM’s improper reasoning about dates, leading to several false positives where the LLM recognized myocardial infractions outside of the past six months. The same improper temporal reasoning led to poor performance in DIETSUPP-2MOS, where the LLM categorized patients as meeting the criteria based on dietary supplements they had taken at one point but not within the past two months. For ADVANCED-CAD, the issue was much more nuanced; the LLM correctly identified certain relevant information as per the guidance provided, but the patient did not completely fit all the criteria.

Among all criteria, we observed two prominent types of failures. First, several false negatives were attributed to insufficient document retrieval, where the LLM did not correctly classify a patient as meeting a criterion because no relevant context was provided. Finally, when using GPT-3.5 Turbo, we observed cases of hallucination, with the LLM citing evidence that was either not present or found in the prompt rather than the patient documents. Such hallucinations were observed at a far lower rate in results using GPT-4. For detailed instances of each type of error, see Appendix A.

**Table 3:**
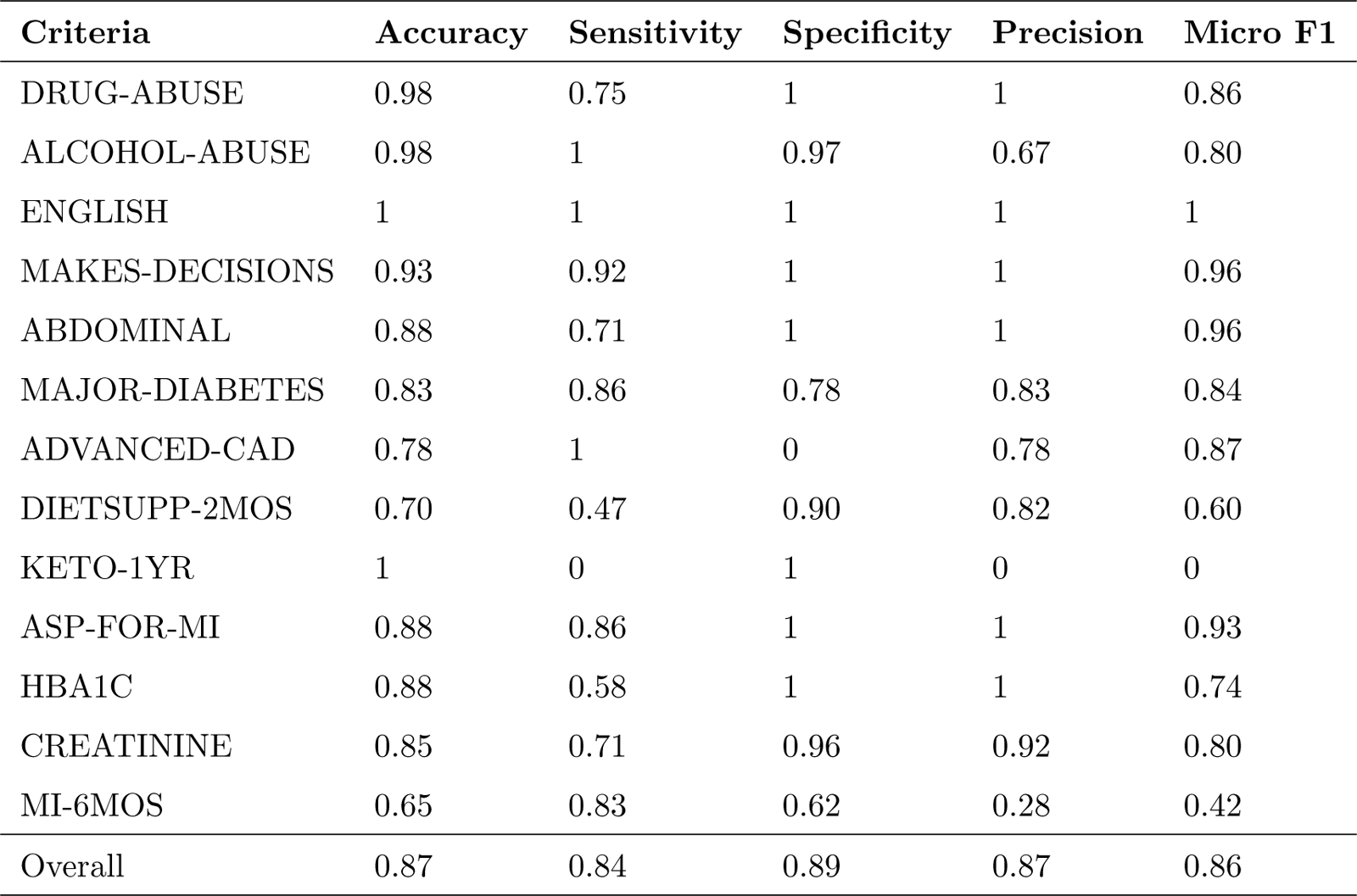
GPT-4 results when utilizing structured JSON output and manually-created expert guidance.

| Criteria | Accuracy | Sensitivity | Specificity | Precision | Micro F1 |
| --- | --- | --- | --- | --- | --- |
| DRUG-ABUSE | 0.98 | 0.75 | 1 | 1 | 0.86 |
| ALCOHOL-ABUSE | 0.98 | 1 | 0.97 | 0.67 | 0.80 |
| ENGLISH | 1 | 1 | 1 | 1 | 1 |
| MAKES-DECISIONS | 0.93 | 0.92 | 1 | 1 | 0.96 |
| ABDOMINAL | 0.88 | 0.71 | 1 | 1 | 0.96 |
| MAJOR-DIABETES | 0.83 | 0.86 | 0.78 | 0.83 | 0.84 |
| ADVANCED-CAD | 0.78 | 1 | 0 | 0.78 | 0.87 |
| DIETSUPP-2MOS | 0.70 | 0.47 | 0.90 | 0.82 | 0.60 |
| KETO-1YR | 1 | 0 | 1 | 0 | 0 |
| ASP-FOR-MI | 0.88 | 0.86 | 1 | 1 | 0.93 |
| HBA1C | 0.88 | 0.58 | 1 | 1 | 0.74 |
| CREATININE | 0.85 | 0.71 | 0.96 | 0.92 | 0.80 |
| MI-6MOS | 0.65 | 0.83 | 0.62 | 0.28 | 0.42 |
| Overall | 0.87 | 0.84 | 0.89 | 0.87 | 0.86 |

## 4 Discussion

In this study, we have shown that OpenAI’s GPT-3.5 Turbo and GPT-4 offer remarkable zero-shot capabilities for clinical trial eligibility screening, achieving an accuracy of 0.81 and 0.87, respectively, both using the structured output and manually-created expert guidance. Our methodology stands out for its user-friendly approach and efficiency, significantly enhancing both aspects compared to traditional screening methods [3, 8]. This improvement is marked by an innovative blend of information retrieval and prompting techniques, allowing clinical research staff to utilize natural language for screening criteria fully. This method not only simplifies the screening process, which is predominantly manual in many clinical settings [4], but also drastically reduces the time required for screening from hours to less than a minute per patient, offering a substantial efficiency gain.

In the 2018 n2c2 challenge, the Medical University of Graz achieved top results with a micro F1 score of 0.91 [14], using a rule-based system reliant on regular expressions and textual markers [9]. The University of Michigan utilized pattern-based, knowledge-intensive methods to achieve a micro F1 score as high as 0.91 [16], placing second in the original 2018 challenge [14]. However, these methods require significant manual effort, with manual annotation and analysis of 202 patient records enabling such high performance. Furthermore, these approaches necessitate complex technical tools for analyzing natural language, posing a barrier to clinical use. These requirements dramatically limit the accessibility and adaptability of this approach. In contrast, our GPT-3.5 Turbo and GPT-4 approaches offer a more user-friendly and flexible solution capable of interpreting patient data in any supported language without extensive setup or maintenance. Utilizing 10% of available patient records, little to no manual analysis, and little to no technical knowledge from end-users, we achieved competitive results. This adaptability significantly broadens the approach’s application and eases the workload for clinical research teams, highlighting the LLMs’ potential to make clinical trial screening more efficient and inclusive.

Our study marks a significant advancement in applying LLMs, particularly GPT-3.5 Turbo and GPT-4, directly facilitating automatic eligibility screening for clinical trials. This approach represents a departure from existing methods, which either rely on rule-based systems with inherent limitations in flexibility and user-friendliness or necessitate manual preprocessing and struggle to interpret complex semantic relationships in clinical data [3, 8]. Unlike previous efforts that utilized LLMs to generate supplemental descriptions for NLP-based models [21], our method leverages LLMs’ advanced semantic analysis capabilities to process unstructured clinical notes directly. This eliminates the need for manual rule definition, preprocessing steps, and additional model training, significantly reducing the operational burden on clinical research staff.

By employing LLMs as the primary engine for screening, we introduce a solution that is more efficient, adaptable, scalable across different clinical contexts. GPT-3.5 Turbo and GPT-4’s ability to autonomously identify and interpret pertinent information from raw clinical notes streamlines the screening workflow, ensuring contextually aware evaluations without the extensive technical expertise required by traditional methods. This innovation underscores the practical benefits of LLMs in clinical trial screenings, enhancing the process’s efficiency and accuracy while offering a scalable and user-friendly tool for clinical research teams.

Our approach, while advancing the use of LLMs in clinical trial eligibility screening, has several limitations, including variability in performance across different criteria, particularly with those that are time-sensitive or necessitate multiple requirements to be met. Additionally, the propensity of LLMs to generate incorrect evidence, known as hallucinations, poses a challenge to achieving consistent and reliable results. These issues highlight the need for further refinement to enhance trust and applicability in clinical settings. To address these challenges, we propose several avenues for improvement. Enhanced prompt engineering techniques, including the adoption of few-shot prompting, offer promising paths to bolster LLM performance directly [17, 2, 19]. Furthermore, leveraging these prompt engineering techniques when creating LLM-generated expert guidance could potentially increase the screening process’s accuracy. Experimenting with sampling answers from the LLM to ensure decision certainty and expanding our method’s testing across diverse clinical datasets will also be critical in identifying and mitigating weaknesses [18]. Finally, recent developments provide a potential path for customizing prompts on a per-criteria basis automatically, further increasing performance and ease-of-use [22]. As we refine our approach, our goal remains to balance accuracy with usability, providing clinical research teams with powerful, easy-to-implement tools to revolutionize patient screening processes.

## 5 Conclusions

In our study, we leveraged the advanced capabilities of GPT-3.5 Turbo and GPT-4, combined with document retrieval technologies, to innovate patient eligibility screening for clinical trials. This approach, utilizing dynamically generated prompts based on expert guidance and raw clinical data, significantly minimizes manual intervention while offering extensive adaptability across medical disciplines. With its promise for scalability and ease of implementation, our method opens new avenues for enhancing the efficiency and effectiveness of clinical trial screenings.

## Data Availability

All data utilized for fine-tuning and testing are available through Harvard University's DBMI Data Portal. All data produced in the present work are contained in the manuscript.

https://portal.dbmi.hms.harvard.edu/projects/n2c2-nlp/

## A Prompting

### A.1 Structured Output

To induce a structured output, we use Pydantic, a data validation library for Python. This allows us to generate a set of formatting instructions automatically. This guides the LLM to produce its output as a JSON object, allowing for easy parsing. As the criteria description, the description and name provided in the original n2c2 challenge were used without modification. The full prompt structure can be seen in **Figure 4**. The output will be provided as a JSON object with keys “criteria name”, “support”, and “criteria”, representing the criteria label, cited evidence, and met/not met status respectively.

**Figure 4:**
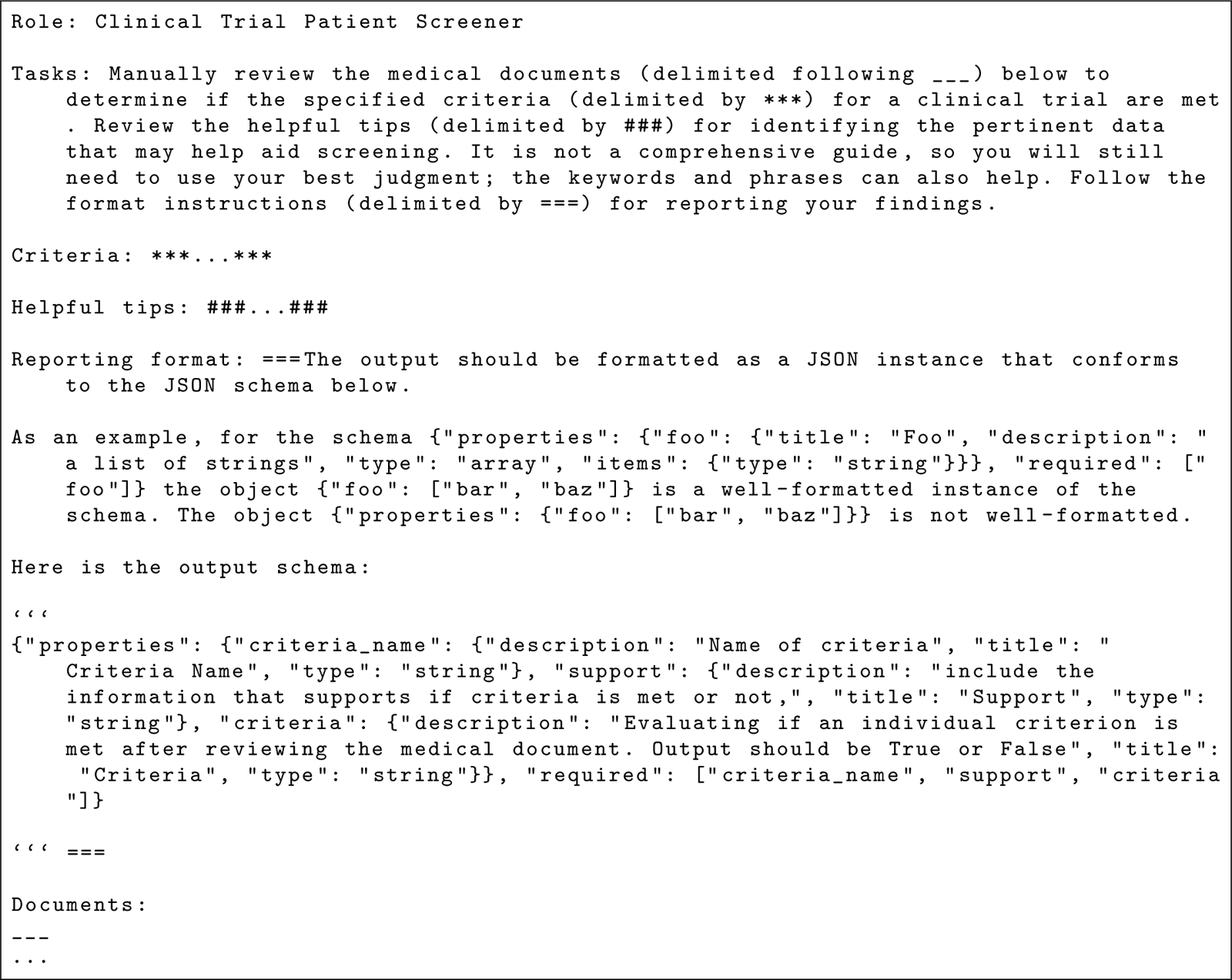
Example prompt for producing structured JSON output

### A.2 Chain-of-thought (CoT) Output

Due to the potential accuracy benefits of inducing CoT reasoning from a LLM, we use a prompt structure that utilizes CoT reasoning. The LLM is prompted to decide, think step-by-step, and provide its thought process. Then, the LLM is asked to use this reasoning to categorize a criterion as “met” or “not met.” In this prompt structure, the n2c2 criteria titles are not included, only the criteria description. The full, prompt structure for ASP-FOR-MI can be seen in **Figure 5**. The output is not structured naturally but is saved into a JSON object with the same format as the structured prompt results for easy analysis.

**Figure 5:**
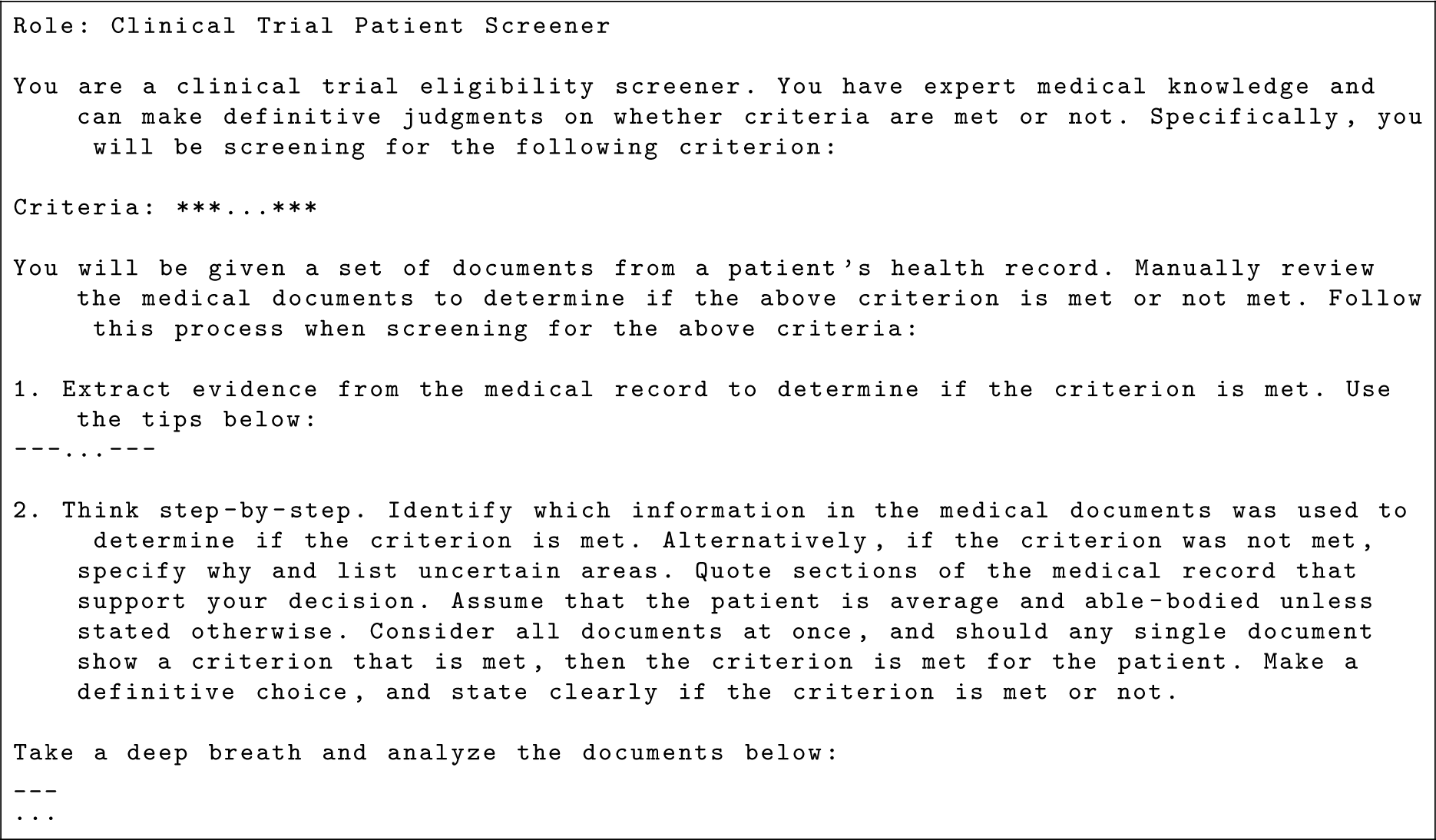
Example prompt for producing CoT reasoning

### A.3 Manual Expert Guidance

Through fine-tuning over 20 patient records, we created a set of expert tips to assist the LLM in the screening process. These tips are inserted directly into the prompt, guiding how to go about the screening process and relevant keywords and phrases. The full, manually created tips for all criteria are listed below.

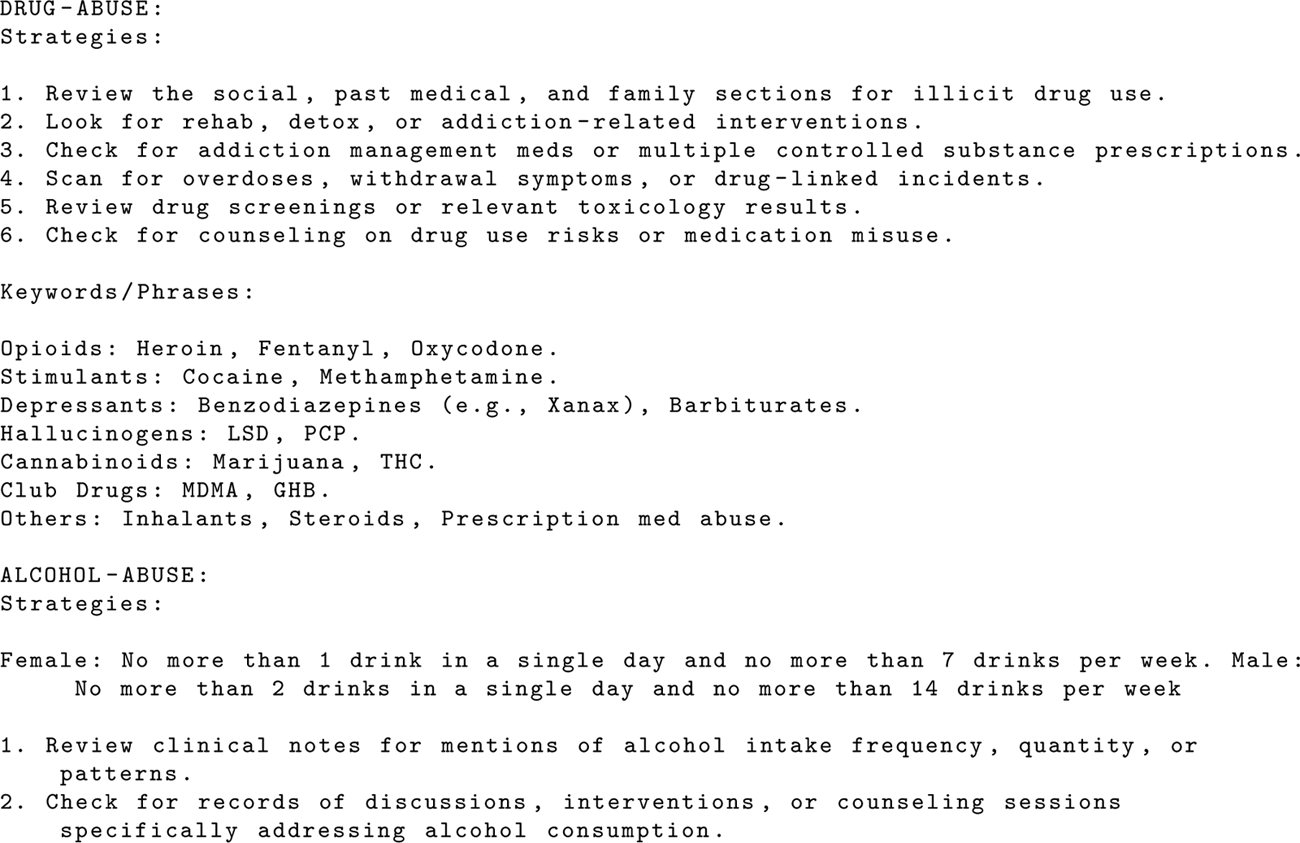
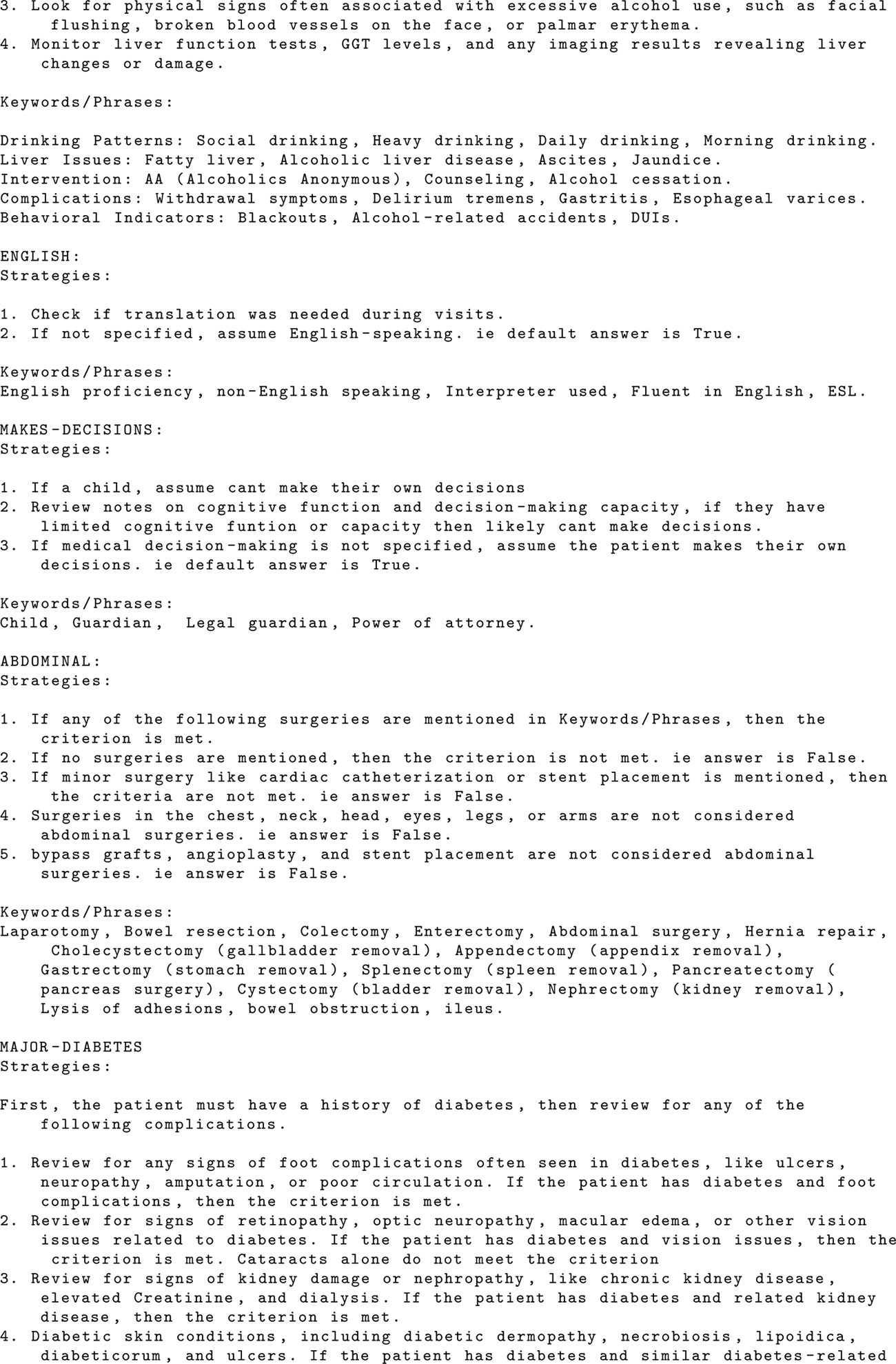
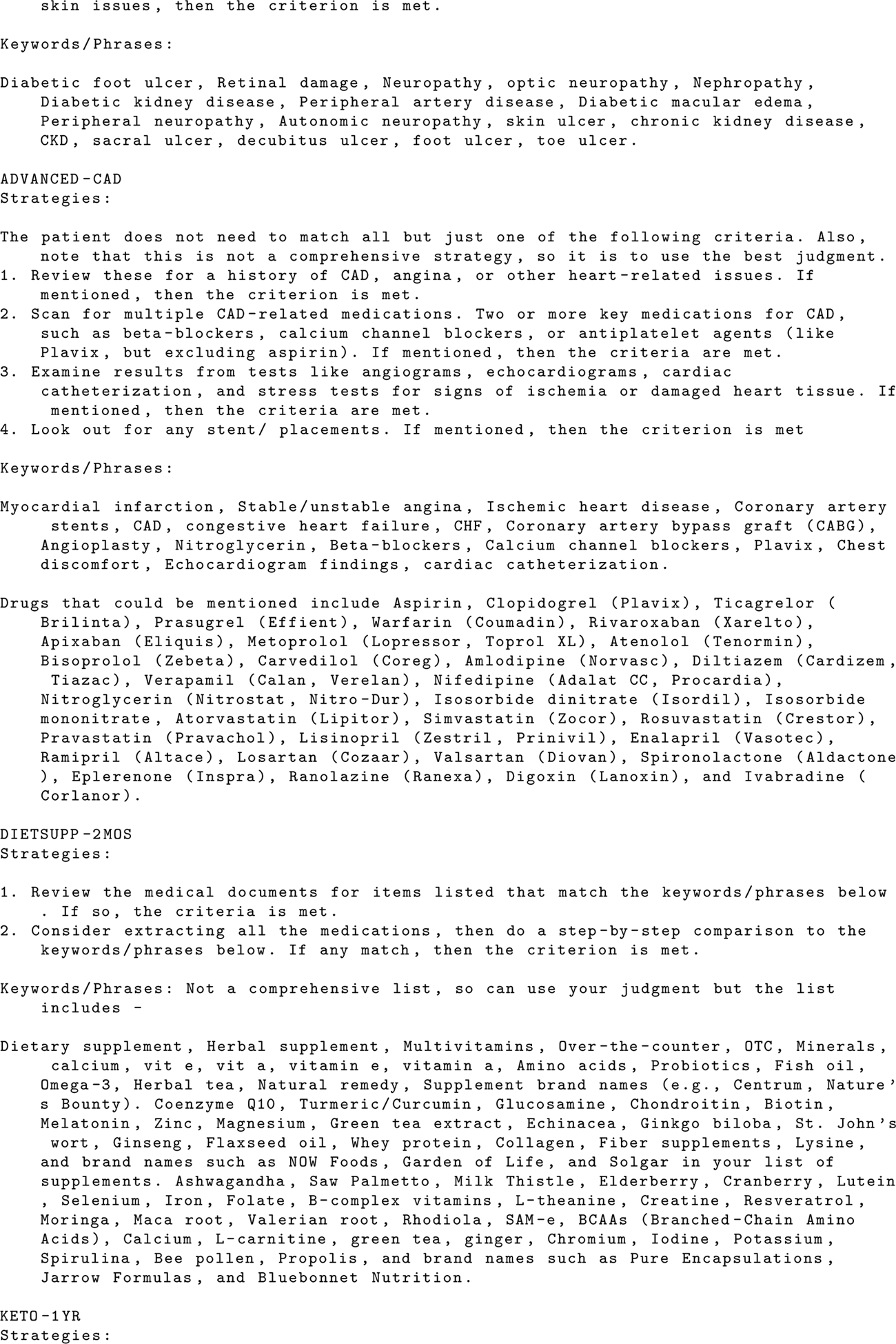
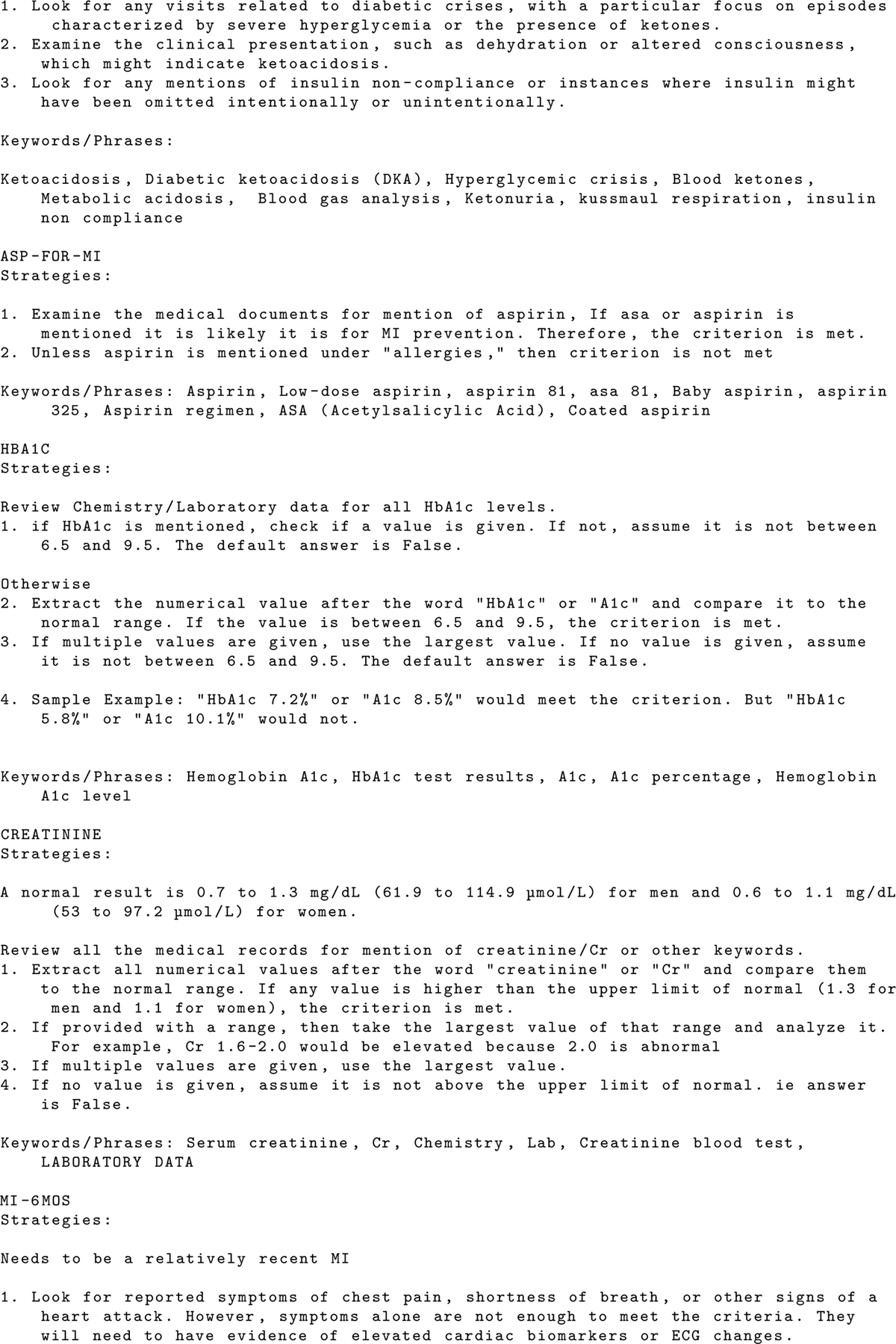
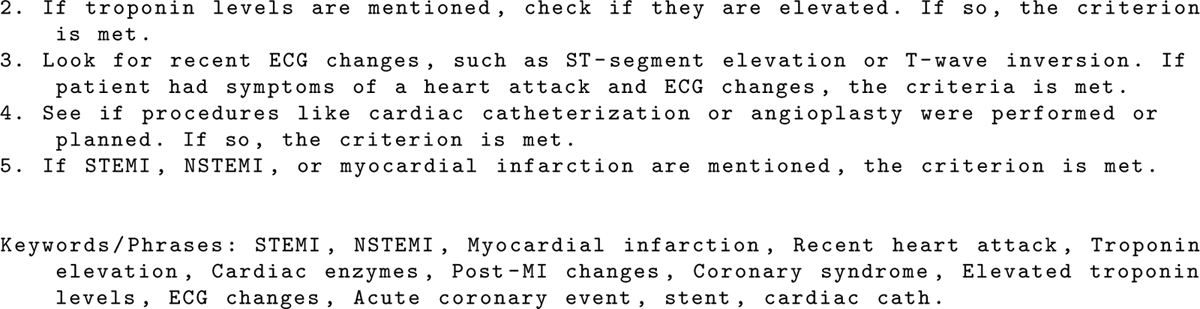

### A.4 LLM-Generated Expert Guidance

To fully automate the eligibility screening process, we used LLMs abilities to generate expert guidance without manual intervention. The LLM is provided with the criteria description and tasked with generating vocabulary, keywords, and phrases associated with the criteria being met or not met. This is done to assist with reasoning and the document retrieval process. The full list of LLM-generated expert tips can be found below.

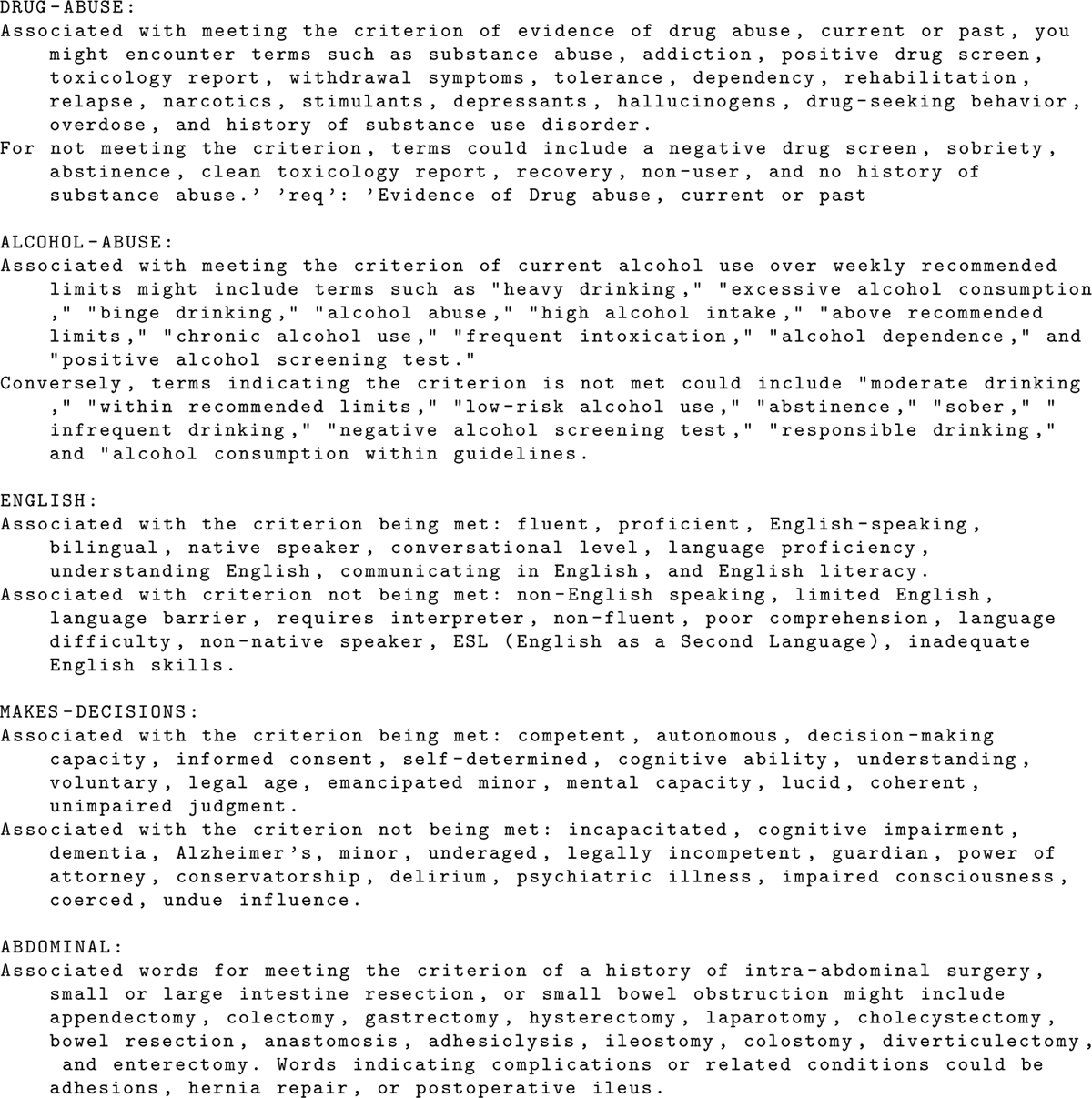
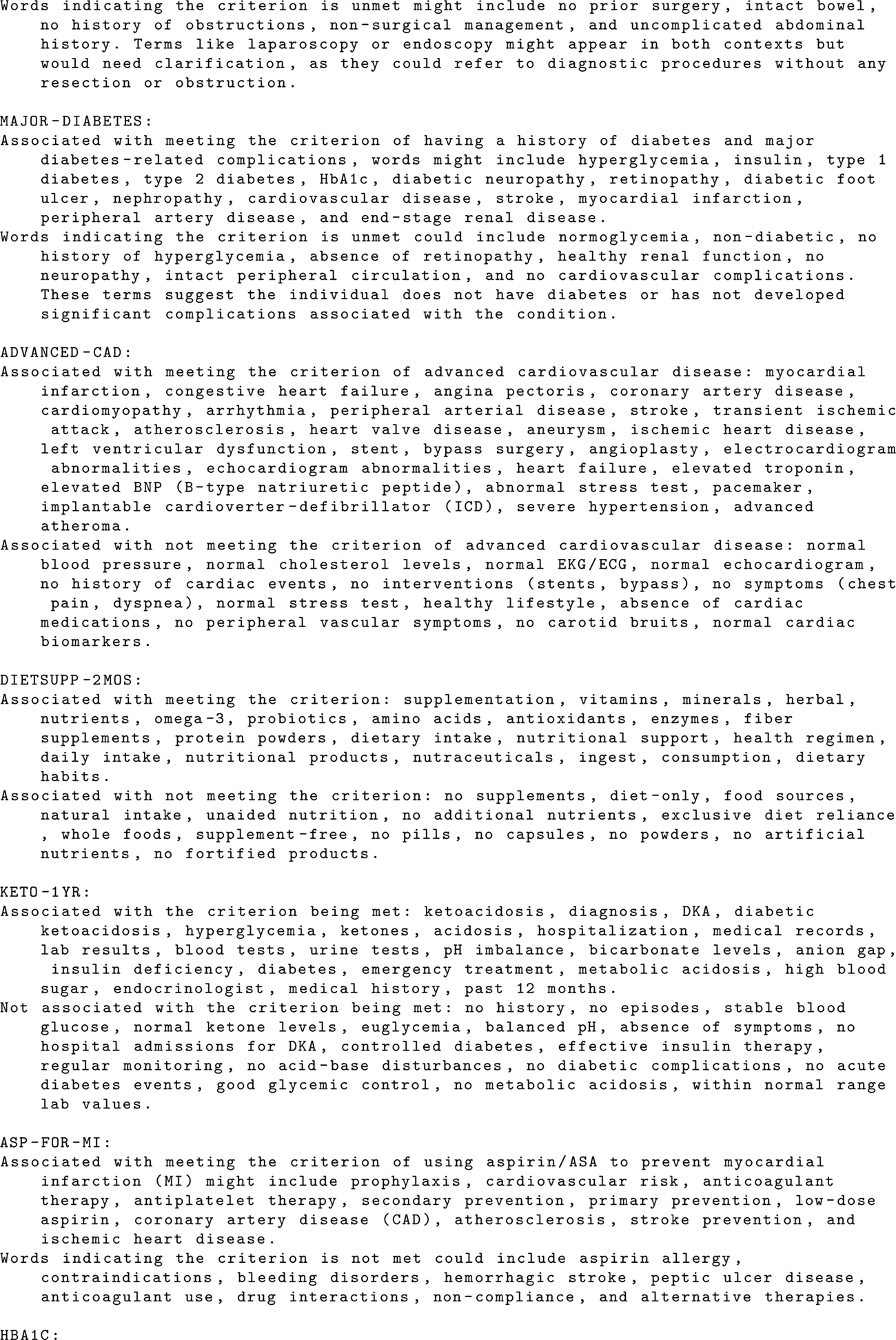
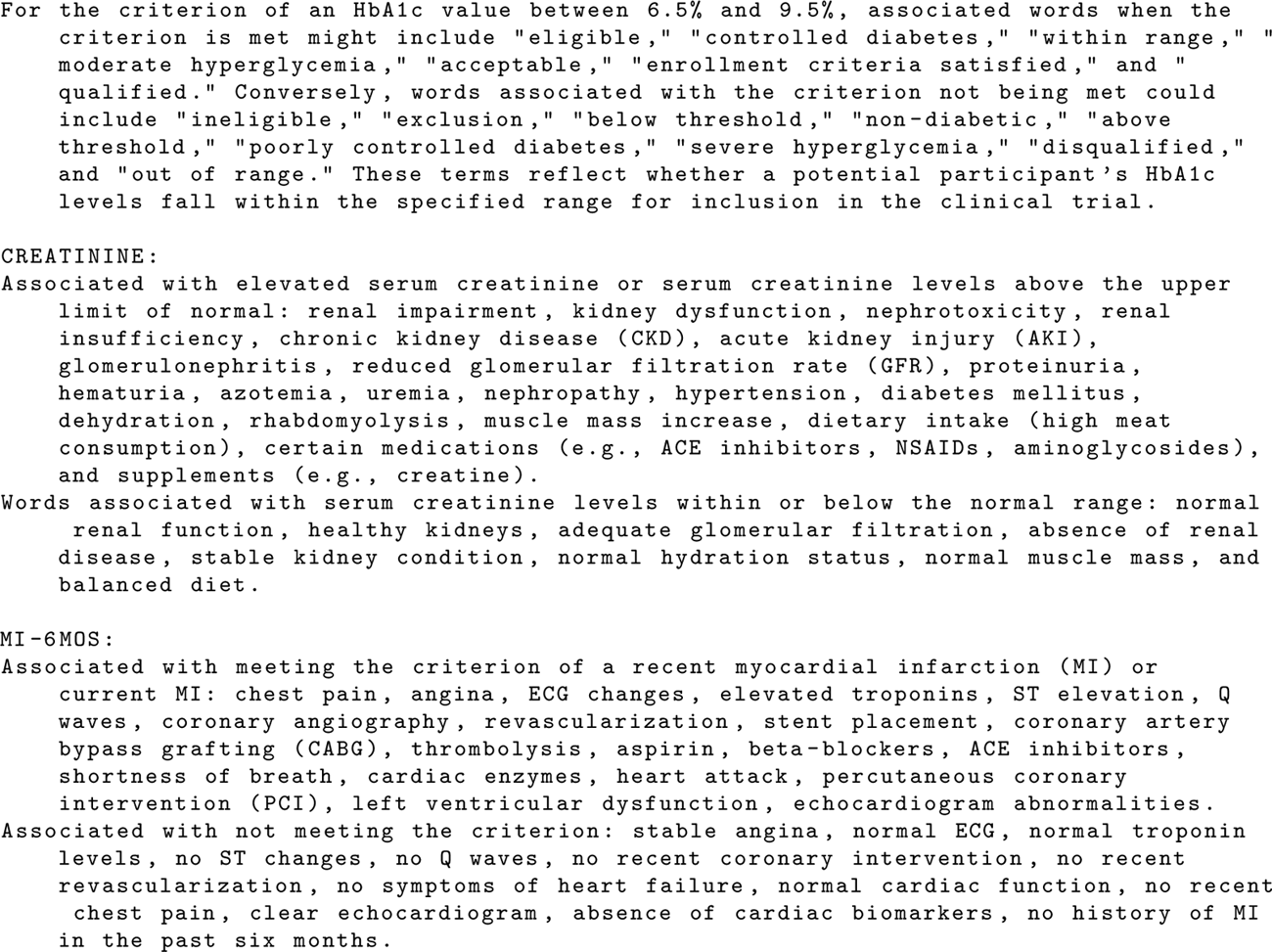

## B Common Failures

Among the results produced by our approach, we observe that incorrect results often fall into one of a handful of categories. With this, the frequency of each type of failure varies based on the criteria type. Specifically, we observe failures due to incorrect logic, uncertainty, or insufficient document retrieval.

### B.1 Incorrect Temporal Reasoning

When screening for date-sensitive criteria, we observed several failures in temporal reasoning. These failures often occurred when evidence was correctly retrieved and identified by the LLM, but a date was not correctly associated with it or reasoned about. For example, when screening a patient for MI-6MOS, the LLM often produces results as shown in **Figure 6**. False negatives due to errors in temporal reasoning were also observed but were much rarer.

**Figure 6:**
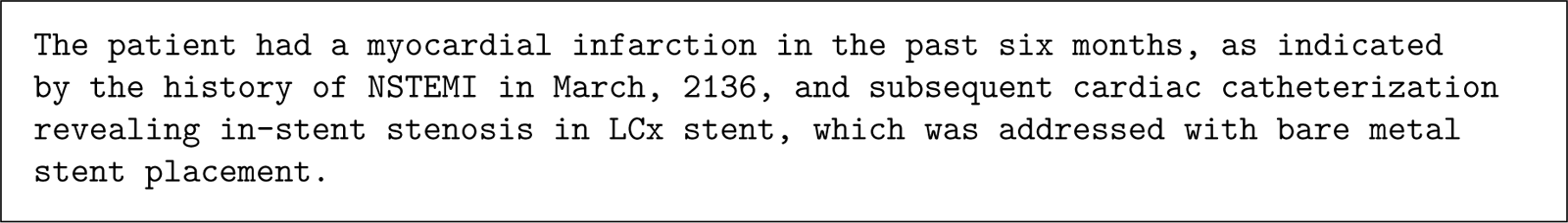
LLM response with incorrect temporal reasoning.

### B.2 Incorrect Logic

In some cases, the LLM correctly identifies evidence but makes an incorrect conclusion based on said evidence. These cases seem most common when screening complex criteria with multiple requirements, such as ADVANCED-CAD or MAJOR-DIABETES. These errors seem to be due to the LLM failing to understand the criteria requirements fully. In the case of ADVANCED-CAD, false positives are common due to the LLM identifying evidence that meets some criteria requirements but not a sufficient amount. An example of this type of error is shown in **Figure 7**, where the LLM classifies a patient as meeting ADVANCED-CAD, but the ground truth rules that the patient does not meet the criteria.

**Figure 7:**
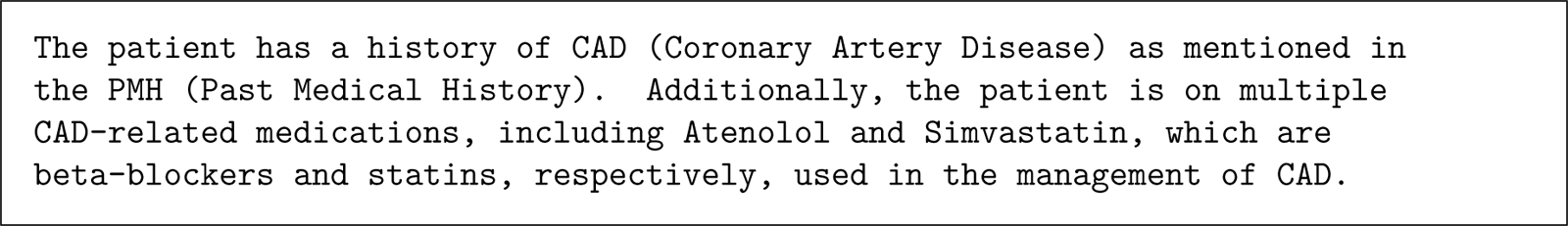
LLM response with incorrect logic.

### B.3 Insufficient Retrieval

Failures across all criteria can be attributed to insufficient document retrieval or information extraction from the LLM. This type of failure is commonly associated with false negatives, as the LLM is not provided with relevant context from the patient record. In most instances of this failure, reasoning such as “No evidence found in the EHR” is listed. This may cause generally lower sensitivity than specificity values across all models.

### B.4 Hallucinations

In our testing, we found that hallucinations of evidence comprised a sizable number of misclassifications when using GPT-3.5 Turbo and a much smaller number of misclassifications when using GPT-4. The LLM often cited evidence from the prompt or expert guidance, even when this information is not found in the patient documents. For example, several patients with varying HbA1c values were incorrectly classified as meeting the HBA1C criterion. The LLM often cited “HbA1c 7.2%” as evidence in these cases, reflecting a portion of the manually-created expert guidance.

